# Testing tongue swab samples by Cepheid Xpert MTB/RIF Ultra: Comparison of two protocols applied to samples from persons with low-bacillary load TB

**DOI:** 10.1101/2025.05.14.25327613

**Authors:** Rachel C. Wood, Sophie Goodgion, Alaina M. Olson, Angelique K. Luabeya, Mark Hatherill, Gerard A. Cangelosi

**Affiliations:** Department of Environmental & Occupational Health Sciences, University of Washington, Seattle, United States of America; South African Tuberculosis Vaccine Initiative, Institute of Infectious Disease & Molecular Medicine and Department of Pathology, University of Cape Town, Cape Town, South Africa

## Abstract

Tongue swabs (TS) are novel non-sputum specimens for molecular diagnosis of tuberculosis (TB). Analysis of swabs by using Cepheid Xpert MTB/RIF Ultra® (Xpert Ultra) is less sensitive than the use of certain manual qPCR methods but is desirable given the widespread use and familiarity of Xpert Ultra. This study evaluated an easy-to-use protocol for analyzing TS by Xpert Ultra. TS samples from symptomatic South African patients were previously collected and tested by Xpert Ultra using a protocol that requires a heating block (termed Heat+TE). Replicate, paired samples from the same patients were tested by a newly reported method that does not require heating (termed 2:1 SR). The diagnostic accuracy of the two methods was compared. Because the sensitivity of TS testing is lowest in patients with low sputum bacillary loads, the resolving power of this paired comparison was maximized by prioritizing samples from such participants (N=108), 88 of whom were TB-positive by sputum microbiological reference standard (MRS) and 20 were TB-negative. Within this sample set, the Heat+TE method was 42.5% (38/88; 95CI 33-54%) sensitive and the 2:1 SR method was 52.3% (46/88; 95CI 41-63%) sensitive relative to sputum MRS. Both methods were 100% (20/20; 95% CI 83-100%) specific. A secondary analysis compared the sensitivities of single-sample versus dual-sample testing within a cohort (N=241) that included both high- and low-bacillary load participants. Testing of a single sample was 75.5% (182/241; 95CI 70-81%) sensitive relative to sputum MRS, whereas dual sampling was 81.3% (196/241; 95CI 76-86%) sensitive.

## Introduction

Pulmonary TB is usually detected by analysis of patient sputum, a mixture of saliva and mucus expectorated from the respiratory tract. Sputum collection presents exposure risks to healthcare workers and many patients cannot routinely produce sputum for testing. Sputum collection can be especially challenging in community settings among minimally- or non-symptomatic people who may comprise the majority of TB cases. These challenges have created strong incentives to identify alternative patient samples that are easier to collect from any human, in any setting (1–4).

Some of these needs can be met by tongue swabbing in combination with nucleic acid amplification testing (NAAT) for *Mycobacterium tuberculosis* (MTB) DNA (5–15). Tongue swabbing is fast, painless, and does not require infrastructure for privacy or infectious aerosol control. Modeling, user preference, and feasibility studies have indicated that tongue swabbing is preferred by patients and health care providers and has the potential to increase the coverage and yield of TB testing, especially in community settings (16–19).

Much ongoing research has focused on identifying the best read-out methods for testing tongue swabs (TS) for MTB DNA. There is evidence that the most sensitive approaches are NAAT methodologies that were developed explicitly for TS samples (5, 14, 15, 20). In contrast, sputum testing platforms such as Cepheid GeneXpert MTB/RIF Ultra® (Xpert Ultra) have exhibited somewhat reduced sensitivity with TS, typically 72% to 77% relative to sputum microbiological reference standard (sputum MRS) (7, 8, 13). However, Xpert Ultra has the advantage of being widely available in TB clinical settings and familiar to users. Therefore, there is considerable interest in adapting Xpert Ultra to the task of TS testing.

In recent studies, we and others tested TS samples by Xpert Ultra using a sample processing method termed “Heat+TE” (7, 13). The Heat+TE method involves first heating the samples to 100 °C for 10 minutes, followed by two applications of 1 mL TE Buffer, vortexing to mix, and transferring all the eluate into the Xpert cartridge. A limitation of this method is that the first step requires a heating apparatus, which is not commonly available at many sites that use Xpert Ultra for sputum testing. Additionally, some sites that applied this method to swabs saw pressure errors rates as high as 16% (13). More recently, a modified swab processing protocol was reported that does not require heating and instead uses a variation of the Cepheid Sample Reagent (SR) protocol that is routinely used for sputum testing on Xpert Ultra (21). An additional advantage of the new method, termed the “2:1 SR” protocol, is that it is reported to cause fewer overpressure errors on the Xpert Ultra platform than the Heat+TE protocol (21). The 2:1 SR method was reported to have a lower (better) limit of detection than the Heat+TE method in laboratory assessments using contrived samples (21). However, the two methods were not directly compared to each other in head-to-head assessments using clinical samples from people with possible TB. The current study addresses that question.

Regardless of analytical method used, numerous studies have shown that people with high bacillary loads in their sputum (e.g., sputum Xpert Ultra semi-quantitative values of High or Medium) are consistently positive by TS regardless of the analytical methodology used (5, 7, 8, 14, 15). Therefore, in order to maximize the resolving power of the current comparison of two analytical methods, we focused on participants who had low sputum bacillary loads or were false negative by the Heat+TE method.

In a secondary analysis, we asked whether the diagnostic yield of tongue swab testing is increased by testing multiple samples from the same individual. During the COVID-19 pandemic, it was common for symptomatic people to exhibit variable COVID-19 test results over the course of their disease (22). We have observed the same with TB oral swabbing. Our first study (12) collected cheek swabs from South African participants with possible TB on 3 consecutive days. Aggregate sensitivity relative to sputum MRS (swab-positive on at least 1 of the 3 days) was 90% (18/20 participants). However, single-day sensitivities were lower, ranging from 60% to 80% (12/20 to 16/20). Our subsequent study (10) tested 2 TS samples collected from participants with possible TB over 2 days. Aggregate sensitivity relative to sputum microbiological reference standard (MRS) was 83% (49/59) while single-day sensitivities were 71% (42/59) and 78% (46/59) on Day 1 and Day 2, respectively. As with SARS-CoV-2 detection in nasal swabs, MTB pathogen loads in oral swabs may vary from day to day, such that diagnostic yield can be increased by serial sampling and testing. A limitation of past studies on serial tongue swab sampling was the use of manual qPCR methods that are very sensitive but not practical for routine use. Xpert Ultra is more practical for routine use, however there is a risk that repeat testing with Xpert Ultra will not exhibit the improved diagnostic yield observed with higher-sensitivity methods. Therefore, the current study evaluated the impact of repeat tongue swab testing using Xpert Ultra as the readout.

## Methods

### Study population

This study used banked TS samples (regular Copan FLOQswabs (520CS01; Copan Italia, Brescia, Italy)) that had been previously collected from South African participants. These participants were enrolled and sampled by the University of Cape Town (UCT) and the South African Tuberculosis Vaccine Initiative (SATVI) (7). The University of Washington (UW) Human Subjects Division (STUDY00001840) and the UCT Human Research Ethics Committee (reference number 160/2020) approved this project.

The previous study (7) collected swab samples from two cohorts of volunteers. In Cohort 1, all participants were TB-positive by a sputum MRS. Sputum MRS positivity was defined as a positive result by sputum Xpert Ultra and/or sputum culture. In Cohort 2, participants were recruited and sampled based on symptom criteria and thus comprised both confirmed TB-positive participants and confirmed TB-negative participants. TB positivity in Cohort 2 was confirmed by sputum MRS as in Cohort 1.

### Heat+TE protocol

In the previous study, TS from Cohort 1 and Cohort 2 participants were tested by Xpert Ultra using a method that involved heating the swab samples to 100 °C, adding TE buffer, vortexing, and then pipetting the sample solution into an Xpert Ultra cartridge (the “Heat+TE” method) (7). Since multiple samples were collected per participant, there were extra samples from most participants that were archived for use in subsequent studies, including the current one.

### 2:1 SR protocol

The 2:1 SR method used in this analysis was reported by others (21) and proceeded as follows. First, diluted SR was prepared by mixing 8 mL of Cepheid SR buffer and 4 mL of 1x PBS buffer. This solution was vortexed at max speed (Vortex Genie 2, VWR) for 5-10 s and used for same-day testing. Swab samples were removed from the −80 °C freezer and thawed on ice for 15 minutes. Working in a biosafety cabinet, 700 μL of the 2:1 diluted SR was added to each sample. The samples were then vortexed at max speed for 5-10 s, followed by an incubation at room temperature (22°C) for 15 minutes. Mid-way through the incubation, between 7-8 min, the samples were vortexed again at max speed for 5-10 s. During the incubation, Xpert Ultra cartridges were labeled with the appropriate sample ID and prefilled with 1.5 mL of 1x PBS buffer. After the incubation, the samples were briefly pulse-vortexed, and 500 μL of the sample fluid from the tube containing the swab and 2:1 diluted SR was added to the Xpert Ultra cartridge. The sample fluid was dispensed into the cartridge slowly against the interior wall to avoid generating bubbles. The cartridge was then inserted into the GeneXpert machine and run according to the manufacturer’s instructions.

### Selection of samples for comparison of methods

Sample selection for this analysis is outlined in **Figure 1**. Samples were chosen from participants who already had a sample tested by Heat+TE (7). To maximize the rigor and resolving power of the paired comparison between Heat+TE and 2:1 SR, samples from participants with low bacillary loads were prioritized. These were identified in two ways. First, we identified participants (N=38) who were true-positive by Heat+TE in the previous study (7), had low sputum semi-quantitative values by Xpert Ultra testing of sputum, and had banked samples available for testing in the current study. Sputum semi-quantitative values were known only for a subset of Cohort 2 participants, therefore in the current study these samples all came from Cohort 2. Of the 38 samples selected for this study, 30 (79%) were from participants with Low, Very Low, or Trace values by sputum Xpert Ultra. By comparison, in the original cohort 32/91 (35%) had sputum semi-quantitative values in these categories (7).

**Figure 1.**
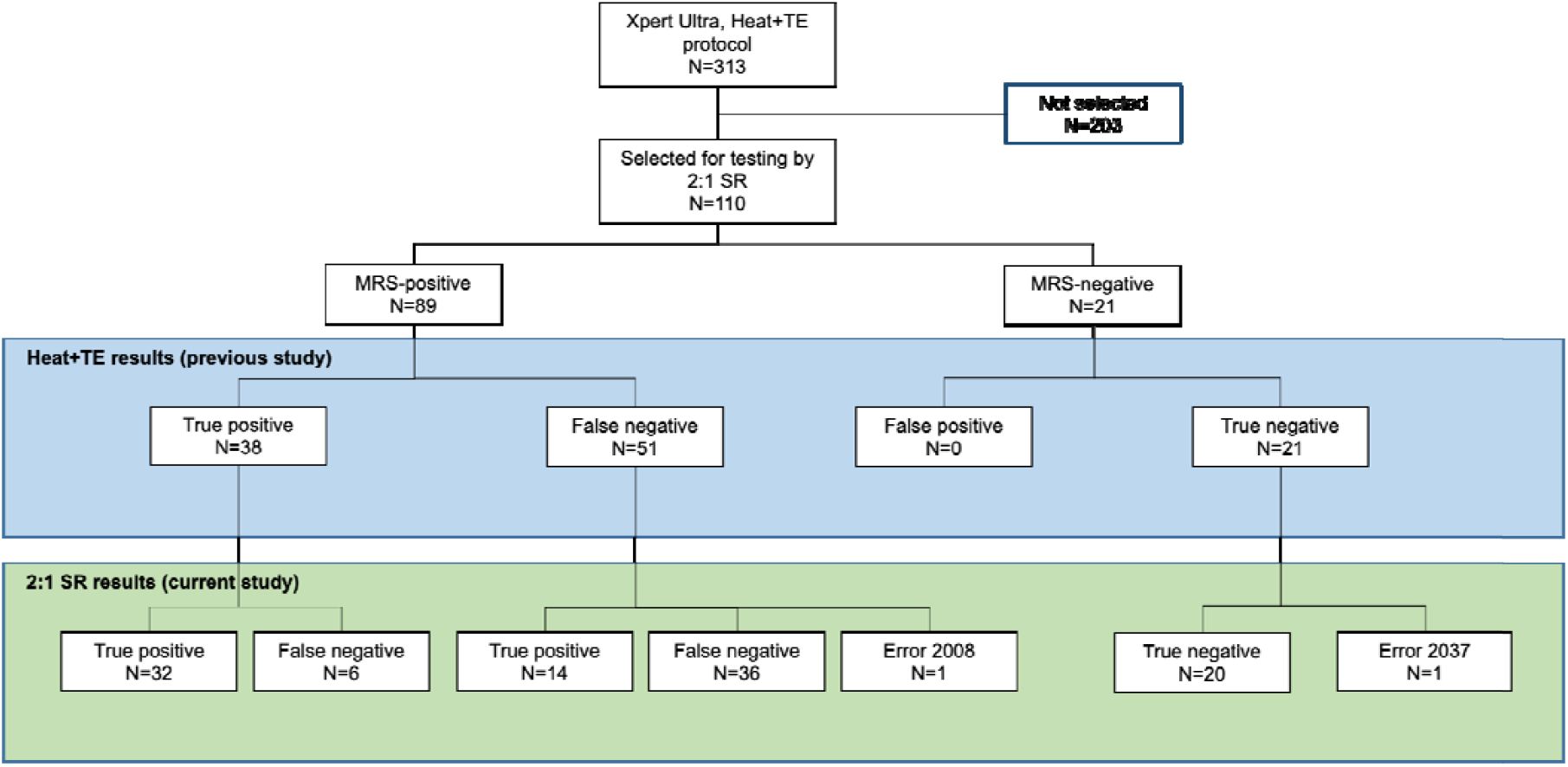
Flowchart displaying the selection of samples tested previously by the Heat+TE method, and the results of testing by the 2:1 SR method.

We also tested samples from all participants who were previously sputum MRS-positive and Heat+TE negative (false-negative by TS) (7). We tested all samples from such participants who had a banked sample available for testing (N=26 from Cohort 1, and N=25 from Cohort 2; total N=51).

Because three consecutive tongue swab samples had been collected from each participant in Cohort 1, the three samples were randomized by collection order so that each swab (first, second, or third) was equally represented. In Cohort 2, five Copan tongue swabs had been collected on Day 1 and two more had been collected on Day 2. The first two samples from Day 1 and the two from Day 2 were included in the previous study (7). For the current study, any of the 5 swabs from Day 1 and either of the two from Day 2 were eligible for inclusion. Samples were randomized by collection day and collection order. To do this, each sample was assigned a number and an online random number generator (randomlists.com) was used to make a list of samples.

To assess the specificity of the 2:1 SR method, leftover samples from 21 sputum MRS-negative participants were tested (**Figure 1**). These were also randomized by collection day and collection order using the online random number generator.

### Assessment of dual sampling

To assess whether dual sampling boosted aggregate sensitivity, the results of all samples tested by Heat+TE (N=313), as reported in Wood *et al*. 2024 (7) were considered alongside the 2:1 SR results in this study. For each participant who had samples tested by both Heat+TE and 2:1 SR, any positive result by either test was considered a swab positive.

### Statistical analysis

Microsoft Excel and GraphPad Prism were used for statistical analyses.

## Results

### Participant characteristics

Replicate swabs from 110 of the 313 participants whose samples were previously tested by Heat+TE (7) were selected for testing by the 2:1 SR protocol as described in Methods. The sample set included N=89 samples from patients with sputum MRS-confirmed TB, and N=21 samples from randomly selected sputum MRS-negative participants **(Figure 1)**. The demographic characteristics of these 110 participants are shown in **Table 1**.

**Table 1.** Characteristics of the participants whose samples were tested by the 2:1 SR protocol for Xpert Ultra.

|  | Cohort 1 | Cohort 2 |  |
| --- | --- | --- | --- |
|  | Sputum MRS-positive (N=26) | Sputum MRS-positive (N=63) | Sputum MRS-negative (N=21) |
| Age (median, interquartile range) | 38 (29-48) | 36 (26-47) | 37 (28-48) |
| Gender (male) | 15 (57.7%) | 35 (54%) | 6 (28.6%) |
| Mixed race ancestry | 19 (73.1%) | 51 (79.4%) | 14 (66.7%) |
| Black | 7 (26.9%) | 13 (20.6%) | 6 (28.6%) |
| Asian | 0 | 0 | 1 (4.8%) |
| HIV-positive | 10 (38.5%) | 24 (38.1%) | 4 (19%) |

### Comparison of Heat+TE to 2:1 SR

**Figure 1** summarizes the results. The Heat+TE results in the diagram were generated previously by Wood *et al*. 2024 (7). Testing the samples from 110 participants yielded 108 valid results and 2 Xpert Ultra error results: Error 2008 (pressure error) and Error 2037 (cartridge error).

Among the N=38 participants with true-positive TS results in the previous study, 32 yielded positive results by the 2:1 SR method (**Figure 1**). This left 6 participants whose samples were false-negative by 2:1 SR. The apparently lower sensitivity of 2:1 SR relative to Heat+TE (84% and 100%, respectively) may have been an artifact of the design of this portion of this study, because all samples were Heat+TE true positive upon selection. Therefore, we selected a second group of sputum MRS-positive participants who were Heat+TE false-negative (N=51; **Figure 1**) and applied the 2:1 SR method to their replicate swab samples. One of these samples yielded an error. Of the remaining 50 swabs, 14 yielded true-positive results by 2:1 diluted SR.

Combining the two sputum MRS-positive sample sets (total N=88 excluding the single error), the sensitivities of the two methods are shown in **Table 2**. The sensitivity of 2:1 SR was higher than the Heat+TE method, but the difference was not statistically significant (*p*=0.23, 2-population proportion z-test). Of the 52 true-positives by either method, 32 were positively identified by both Heat+TE and 2:1 SR, while 20 were positively identified by just one or the other method (**Figure 2**).

**Table 2.** Sensitivity and specificity of the Heat+TE and 2:1 SR protocols relative to sputum MRS.

|  | **Sensitivity** | **Specificity** |
| --- | --- | --- |
| Heat+TE | 42.5% (38/88); 95% CI 32.7%, 54.2% | 100% (20/20); 95% CI 83.2%, 100.0% |
| 2:1 SR | 52.3% (46/88); 95% CI 41.4%, 63.0% | 100% (20/20); 95% CI 83.2%, 100.0% |

**Figure 2.**
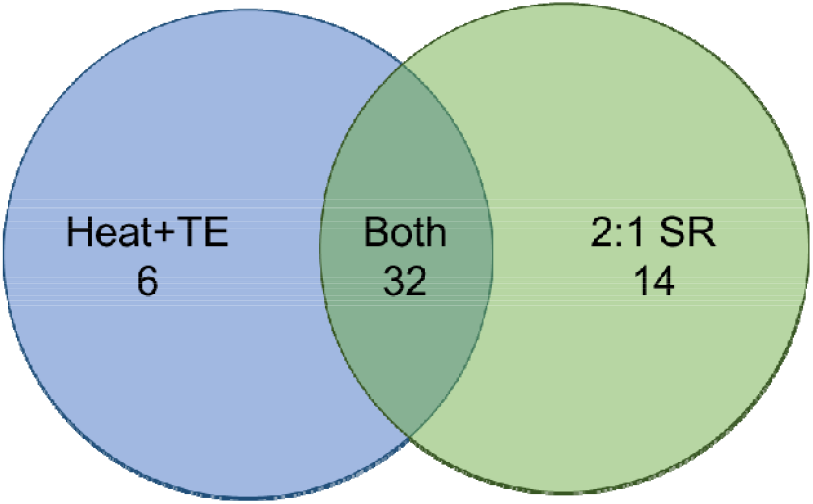
Venn-diagram showing the number of true-positive samples by both methods and either method.

### Impact of dual sample analysis on diagnostic accuracy

Our previous analysis (7) used Heat+TE with Xpert Ultra to test TS samples from 313 South African participants, 241 of whom had sputum MRS-confirmed TB. The sensitivity of the method was 182/241, or 75.5% (95% CI 69.6%, 80.8%). The present study tested a second sample from 51 of the 59 participants who were false-negative by TS in the previous study. Fourteen of these samples yielded positive results, albeit by a modified methodology relative to the previous study. This raised the combined sensitivity of TS (positive on at least 1 swab) to 196/241, or 81.3% (85% CI 75.8%, 86.0%).

### Tongue swabs by Xpert Ultra among people living with HIV

Among the entire cohort from which swabs were tested by the Heat+TE protocol (7), 33% were PLHIV. Among those participants for whom an extra swab was tested in this study by the 2:1 SR protocol, 35% were PLHIV. For both methods, sensitivity was lower among PLHIV than among people without HIV (**Figure 3**).

**Figure 3.**
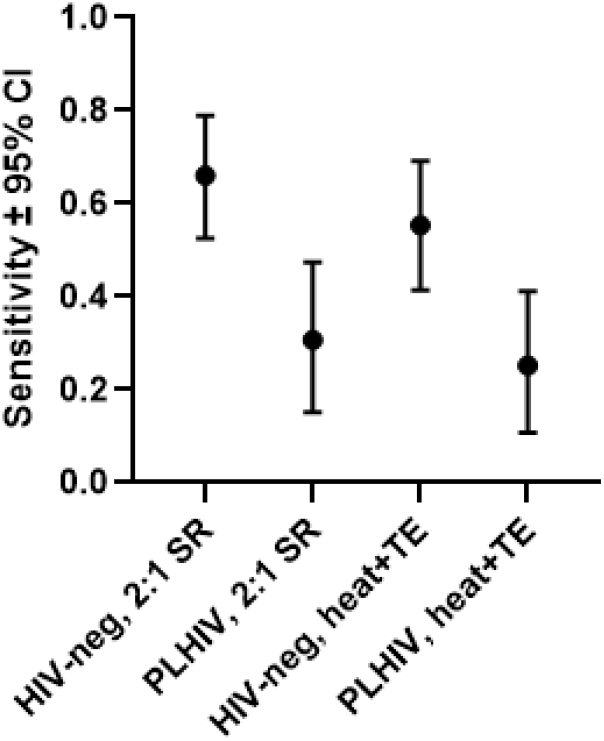
Sensitivities of the two methods relative to HIV status.

## Discussion

The 2:1 SR method was developed to mitigate two limitations of the Heat+TE method when used to prepare swab samples for Xpert Ultra testing, namely the requirement for a heating instrument and the tendency of Heat+TE to cause overpressure errors in some populations. Here, we used a paired sample comparison to show that the advantages of 2:1 SR come at no cost with regard to the rate of positive detection of TB. In fact, the sensitivity of the 2:1 SR method appeared higher than the Heat+TE method, though the two had overlapping confidence intervals. The sensitivities of both methods reported here are artificially low due to the prioritization of samples from low-bacillary load participants. This does not reflect the sensitivity that could be expected if applied to a broader population of symptomatic patients.

High error rates in some populations were reported with the Heat+TE protocol and ranged from 0-16% (13). In this smaller study, the overall error rate of the 2:1 SR method was 1.8% (2/110) and the rate of pressure errors was 0.9%. Given that the initial error rate reported for Heat+TE by Wood *et al*. 2024 (1%) was also low, this study could not assess whether the 2:1 SR method reduced the problem.

However, *in vitro* experiments with contrived samples, reported by Andama *et al*. (8) and Chilambi *et al*. (21), showed that the addition of Cepheid SR to TS samples may reduce error rates in Xpert Ultra.

As has been the case previously (5, 10), the sensitivity of both the Heat+TE method and the 2:1 SR method dropped when used on samples collected from PLHIV. PLHIV tend to have lower bacillary load infections than those without HIV (23), which could be the driver of this difference in swab detection.

The results of the present study showed that the 2:1 SR method is no more or less sensitive to this quantitative effect than Heat+TE.

Xpert Ultra testing of two serially collected samples resulted in only a modest (not statistically significant) increase in sensitivity. However, even a modest increase could be beneficial in populations where tongue swabs are easier and more manageable to collect than sputum. Further work using more sensitive, swab-dedicated methods (5, 14) could uncover further improvements in sensitivity through the use of serially collected samples.

This study had several limitations. The sample size was insufficient to fully compare the diagnostic accuracy of the 2:1 SR method relative to Heat+TE. Limitations on materials (including Xpert Ultra kits which are difficult to procure in the United States) prevented us from assessing specificity more fully. Additionally, because two different methods were employed, this study was not able to fully address the utility of serial swabbing as a method of increasing sensitivity. The increase in sensitivity reported here may be due to serial swabbing, or it may be due to the difference in method.

Despite these limitations, this study supports the adoption of the 2:1 SR method as the protocol of choice for processing tongue swabs for analysis by Xpert Ultra. It has methodological and logistical advantages over the previous Heat+TE method, and no detectable disadvantages with regard to diagnostic accuracy.

## Data Availability

All data produced in the present study are available upon reasonable request to the authors.

## Acknowledgements

The authors are grateful to Adithya Cattamanchi for sharing protocols, and to Loren Rockman and Grant Theron for helping to source reagents. This work was funded by NIH grant R21 AI185407.

